# Current characteristics and outcomes of Cytomegalovirus Reactivation in kidney transplant seropositive recipients in the era of prophylaxis treatment. Lesson from single Moroccan center experience

**DOI:** 10.1101/19001008

**Authors:** Bouchra. Rezzouk, Tarik. Bouattar, Bouchra. Belkadi, Rachid. Razine, Rabia. Bayahia, Naima. Ouzeddoun, Loubna. Benamar, Hakima. Rhou, Najat. Bouihat, Azeddine Ibrahimi, Myriam. Seffar, Hakima. Kabbaj

## Abstract

Despite the use of antiviral prophylaxis, the active cytomegalovirus (CMV) replication is still occurred in the seropositive kidney recipients. The aim of this study was to assess the incidence of CMV reactivation and potential risk factors associated with CMV disease. Data of sixty kidney transplant recipients who had received CMV prophylaxis were obtained between 2013 and 2017. Quantitative nucleic acid amplification testing for CMV viraemia was assessed using Abbott RealT*ime* Polymerase Chain Reaction (PCR). Among the seropositive recipients, cumulative incidence for reactivation was 63%. In patients with quantitative viraemia, the time of active replication was significantly lower compared to those with detectable viraemia (141.5 ± 96.9 *vs* 294.1 ± 112.6 days, *P* < 0.001). During prophylactic treatment, 46.7% of patients with quantifiable viraemia had experienced active replication and none among patients with detectable viraemia (*P*= 0.017). Importantly, symptomatic reactivation was significantly observed in the younger patients with higher peak viraemia compared to those with symptoms free (28.8 ± 5.12 vs. 38.1 ± 12.34 years, *P*= 0.007) and 3.8 ± 1.59 vs. 2.4 ± 0. 79 log_10_IU/ml, *P* = 0.003, respectively). Furthermore, the median duration of viraemia (21.2, vs. 13.4 days, *P*= 0.028) and period of CMV therapy (24.3 vs 12.3 days, *P* <0.001) were significantly longer for this group. In addition, intercurrent infections (75% vs. 23%, *P* = 0.028) and acute rejection (50 % vs 0%, *P* = 0.003) were significantly more frequent in symptomatic reactivation group. In addition, peak viral load was a potential risk factor for development of symptomatic reactivation with odds ratio 3.39, 95%CI=1.21-9.53, *P* = 0.02). In conclusion, CMV reactivation remains serious problem for seropositive recipients who were expected to be on antiviral prophylaxis. Patients with high level of viraemia may be at an increased risk of progression to CMV disease and adverse outcomes.

## Introduction

Human cytomegalovirus (HCMV) is an ubiquitous herpesvirus, which causes the common complication in immunocompromised individuals, including the transplant recipients [1–3]. In the absence of antiviral treatment, CMV disease occurs in up to 60 % of transplant recipients [4].

In patients receiving a kidney transplant, CMV plays a critical role in direct induction of infection syndrome and severity disease [5, 6]. Moreover, it can be responsible indirectly for decreased graft function with increased morbidity such intercurrent infections, cardiovascular disease, malignancies or graft loss and long term mortality [7–10]. Previously, seropositive donor with seronegative recipient were considered as high serologically risk patients of occurrence CMV disease [11,12]. But, the risk is also observed currently in seropositive recipients who are exposed to CMV reactivation or reinfection by novel strain of donor [13]. The virus reactivation from latency are likely to occur in immunosuppressed patients receiving intensive immunosuppressive agents and may have serious clinical effects after kidney transplantation [14,15]. Therefore, antiviral prophylaxis is usually recommended for a short period with three months of treatment by recent consensus guidelines [16]. However, the active CMV replication is still occurred in the D+/R+ population over the first three months of prophylaxis [17,18], and associated with CMV viraemia or with late-onset disease [19–21]. Furthermore, CMV recurrence infections can be developed after the first episode of reactivation in this group [22– 24]. However, Several studies focused for investigation in recipients of various solid organ transplant or in the high-risk kidney patients (D+/R-) [25, 26]. In addition, a majority of the experts proposed for prevention prophylaxis, the use of oral valganciclovir or ganciclovir as antiviral medication for three months [5,14]. But, no recent data is known about the risk of CMV disease after reactivation or recurrence in seropositive patients treated with valaciclovir up to three months post kidney-transplantation.

For the present study, only the recipients with specific Ig G antibodies receiving allograft from CMV seropositive donors were taken into account. Furthermore, immunosuppression is frequently conservative and antiviral prophylaxis is performed between 3 to 6 months for all recipients with oral valaciclovir (VACV) or valganciclovir (VGCV). For these reasons, it is of major importance to identify the clinical outcomes of CMV reactivation and to explore the potential risk factors associated with CMV disease after renal transplantation especially in the recipient seropositive population. The aims of this study were to assess the incidence of CMV reactivation and to determine potential risk factors associated with CMV disease after renal transplantation in our recipient’s seropositive population.

## Materials and Methods

### Study population

For this retrospective study, all renal transplant performed between March 2013 and August 2017 at University hospital Ibn Sina (Morocco) were reviewed using patient’s medical records. Throughout this period, all CMV seropositive (IgG) recipients over 15 years old at transplantation, who received live or deceased donor kidney transplant were included in this analysis. Others selection criteria were summarized in flow chart (Figure 1). The study was approved by the institutional ethical committee in biomedical research of Faculty of Medicine and Pharmacy, Rabat (number 26/18-2017).

**Figure 1.**
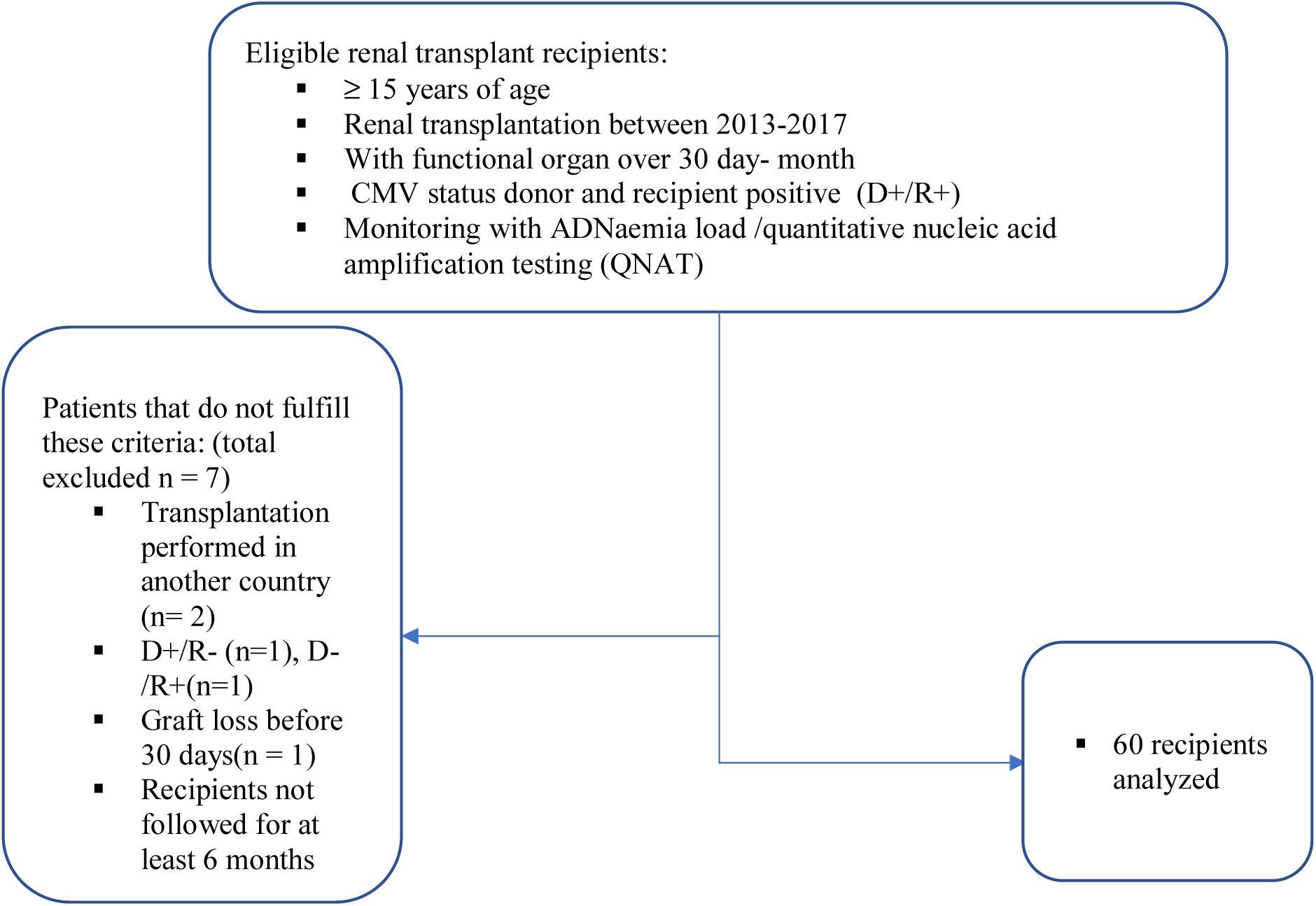
Study Flow chart

### Immunosuppressive regimen and CMV treatment

For the most patients of this study, rabbit antithymocyte globulin (rATG/ Thymoglobuline) has been used as induction therapy and those with high risk of rejection had received basiliximab induction during the first week post transplantation. The induction therapy was given in association with calcineurin inhibitors (tacrolimus or ciclosporin). Generally, immunosuppressive regimen was based usually on tacrolimus (Tac), mycophenolate mofetil (MMF) or mycophenolic acid (Myfortic) and steroids (bolus solumedrol and cortancyl).

Renal graft rejection was diagnosed by renal transplant biopsy and treated with high – dose intravenous corticosteroids and /or switch to tacrolimus if they were initially treated with cyclosporine.

CMV prophylaxis was administered for all patients during three to six months after transplantation with following drugs: oral valaciclovir (VLC) or oral valganciclovir (VGCV) with dosage according to our center practice. Valaciclovir was routinely given at dose 4.5 to 6 **g** once daily and the use of valganciclovir involved for patients with early viral replication at dose 450 mg or 900 mg/ day, dose adjusted according to renal function (estimated glomerular diet of renal disease calculated eGFRD by the Modification of Diet in Renal Disease equation MDRD).

Patients experiencing CMV viraemia without syndrome or disease were classified as asymptomatic viraemia and monitored for CMV DNAemia by RealT*ime* PCR methods until eradication without interventions. If CMV disease was suspected or some patients had high viral load in the absence of clinical signs, a i.v. ganciclovir (250 mg or 500 mg twice daily) was given as main treatment (dose adjusted for renal function). CMV treatment was stopped after CMV DNAemia was undetectable for two consecutive tests. Fewer patients with mild infection or with low viral load had received a valaciclovir (6g/day) or valganciclovir (450-1800 mg/day) as preemptive or curative treatment.

No antiviral treatment was given to patients with kidney function impairment but a reduction of immunosuppression was adopted. Second treatment prophylaxis with valaciclovir was initiated for patients with risk of recurrence.

### Outcomes variables

According to the internationals consensus guidelines and clinical studies definitions [16,27]; CMV infection was defined as “evidence of CMV replication regardless of symptoms (differs from latent CMV), or as detection of viral proteins (antigens), or nucleic acid in any body fluid or tissue specimen”. Asymptomatic viraemia was defined as “presence of quantitative viral load without CMV related clinical symptoms.” CMV disease was categorized as “evidence of CMV infection with attributable symptoms”. CMV disease can be further categorized as “viral syndrome (ie, fever, malaise, leukopenia, and/or thrombocytopenia), or as tissue invasive” [16]. CMV Reactivation was defined as an active viral DNA replication in recipient who had been previously infected by CMV before kidney transplantation. Recurrent cytomegalovirus infection was defined as detectable or measurement of CMV viral load after the first infection episode (achieving initial clearance) [22]. In addition, a detectable viraemia was defined as being the detection of CMV DNA below the quantification threshold (< 1.49 log_10_ IU/ml) using Abbott RealT*im*e CMV assay.

For each patient, Baseline data, follow-up data and data about CMV infections and clinical symptoms were collected from individual patient records. Extracted Baseline data were age recipient at transplant, gender, date of transplantation, previous renal replacement therapy modality (hemodialysis, peritoneal dialysis, or none of patients with preemptive transplantation), primary renal disease, presence of comorbidities, HLA matching (HLA-AB and DR), induction therapy and initial immunosuppressive regimen. For donor, we also recorded demographic characteristics, source of donors (living, deceased), pretransplant CMV serostatus, immunological and serological characteristics.

The follow-up data included liver function test (ALT, AST), immunological follow-up (anti-HLA I and II antibodies), renal function (creatinine and clearance creatine), monitoring of blood count (leukocytes, polynuclear neutrophils, lymphocytes). Data recorded with regard to CMV prophylaxis treatment were the type of antiviral drug (valaciclovir, valganciclovir), doses and duration of prophylaxis treatment. Viraemia data at the time of follow-up included quantification of measurement viral load at diagnosis, the peak viral load, duration of quantifiable viraemia, number of reactivation episodes, duration of onset of replication after transplantation, at the time of prophylactic treatment, after discontinuation of treatment and during posttransplant follow-up. The data of CMV infection included clinical findings as clinicals syndrome with fever, malaise, vomiting, asthenia and headaches or as symptoms disease (diarrhea, colitis, hepatitis, pneumonitis…), Whereas data of immunosuppression treatment during infection covered the types of immunosuppressive drugs and their dosage. Other assessed data included antiviral drugs curative treatment received against CMV (ganciclovir, valganciclovir, and valaciclovir), the length of curative duration and treatment of second prophylaxis (type of drug, length of prophylaxis duration). Occurrence of co-infection as viral infections (BK virus, HBV, HCV, EBV), bacterial infection or fungal infections and acute rejection has been also included.

### CMV assessments by quantitative PCR -CMV

CMV viraemia was assessed every month during CMV prophylaxis, then just one at the time of discontinuation prophylaxis. After the cessation of prophylaxis, patients were monitored once per month during 90 days. All CMV plasma samples were tested at Central Laboratory of Virology using Abbott RealT*ime* CMV assay (Abbott Molecular System Inc., Des Plaines, IL, USA). The review was conducted from March 2013, date of introduction of m2000 Real T*ime* platform which had a linear range of 31.2 to 156 000 000 IU/ml according to the World Health Organization WHO international CMV quantitative standard (National Institute for biological Standards and Control [NIBSC] code 09/162; NIBSC, Hertfordshire, Great Britain) [28]. The threshold of detection was 1.49 log_10_ IU/ml (31.2 IU/ml) and our results were reported in log_10_ IU/ml.

### Statistical analysis

Demographic, clinical, transplant, immunosuppression treatment and antiviral features were described using mean ± standard deviation and /or median for quantitative data. Categorial data were expressed as frequency and proportion. For estimating the timing of CMV reactivation after transplantation, we had used the Kaplan-Meir method. Comparisons in variables distribution between groups were done by using t-student or Mann-Whitney *U*-test for continuous variables and Fischer’s exact test or chi-square test for categorial variables. In addition, univariate logistic regression models were applied for prediction either of CMV symptomatic infection, quantitative viraemia and recurrence. We then selected the most significant variables to define risk factor. The Results from the logistic regression were expressed as odds ratio (OR) and 95% confidence interval (CI). A *p-*value < 0.05 was considered significant and the calculations of data were performed with SPSS v.13 (SPSS, Inc., Chicago, Il, USA).

## Results

### Incidence of CMV reactivation

During the 4-years period (2013-2017), a total of 60 recipients seropositive were analyzed. The mean follow-up after transplantation was 795 ± 444 days [120 - 1580] during which one patient had died (1.7%) and another recipient (1.7%) returned to dialysis. The cause of death was malignancy and the loss of graft was due to primary hyperoxaluria. Most of transplant patients (83%) received thymoglobulin followed by tacrolimus monotherapy (83%). Whereas 10 (17%) were treated only with basiliximab. 38/60 (63%) had developed first episode of reactivation/reinfection after kidney transplantation. The incidence rate of CMV reactivation was 27 per 100 person -year and median time to reactivation was at 227 days [14-1560 days]. Figure 2 depicts the timing of CMV reactivation after kidney transplantation. No differences in the demographics and transplant features were found between the patients with or without CMV reactivation/reinfection (Table 1).

**Table 1a.**
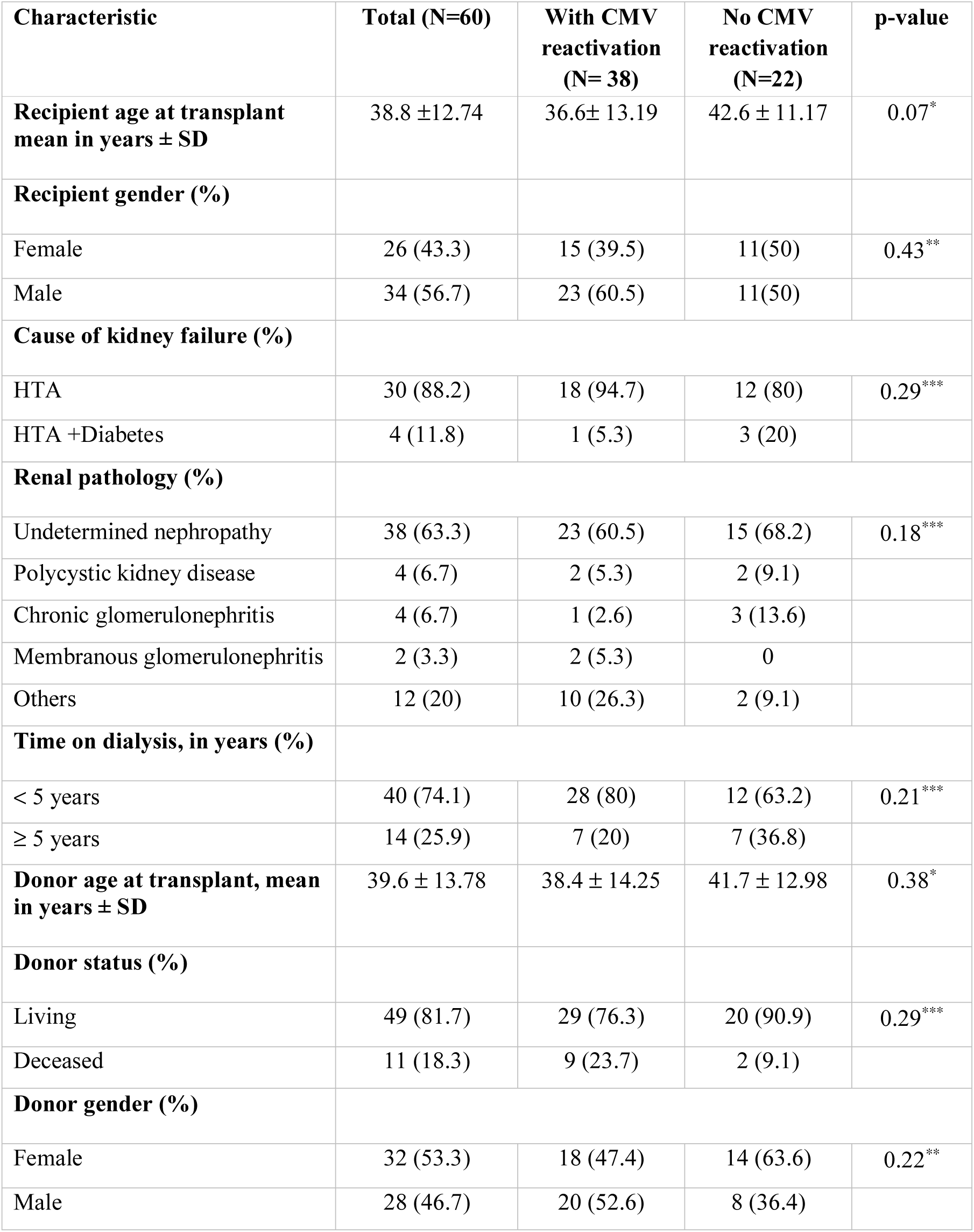
Comparison of patient’s characteristics in those with CMV reactivation and those without, after kidney transplantation.

| Characteristic | Total (N=60) | With CMV reactivation (N= 38) | No CMV reactivation (N=22) | p-value |
| --- | --- | --- | --- | --- |
| **Recipient age at transplant mean in years ± SD** | 38.8 ± 12.74 | 36.6 ± 13.19 | 42.6 ± 11.17 | 0.07* |
| **Recipient gender (%)** |  |  |  |  |
| Female | 26 (43.3) | 15 (39.5) | 11(50) | 0.43** |
| Male | 34 (56.7) | 23 (60.5) | 11(50) |  |
| **Cause of kidney failure (%)** |  |  |  |  |
| HTA | 30 (88.2) | 18 (94.7) | 12 (80) | 0.29*** |
| HTA +Diabetes | 4 (11.8) | 1 (5.3) | 3 (20) |  |
| **Renal pathology (%)** |  |  |  |  |
| Undetermined nephropathy | 38 (63.3) | 23 (60.5) | 15 (68.2) | 0.18*** |
| Polycystic kidney disease | 4 (6.7) | 2 (5.3) | 2 (9.1) |  |
| Chronic glomerulonephritis | 4 (6.7) | 1 (2.6) | 3 (13.6) |  |
| Membranous glomerulonephritis | 2 (3.3) | 2 (5.3) | 0 |  |
| Others | 12 (20) | 10 (26.3) | 2 (9.1) |  |
| **Time on dialysis, in years (%)** |  |  |  |  |
| < 5 years | 40 (74.1) | 28 (80) | 12 (63.2) | 0.21*** |
| ≥ 5 years | 14 (25.9) | 7 (20) | 7 (36.8) | |
| **Donor age at transplant, mean in years ± SD** | 39.6 ± 13.78 | 38.4 ± 14.25 | 41.7 ± 12.98 | 0.38* |
| **Donor status (%)** |  |  |  |  |
| Living | 49 (81.7) | 29 (76.3) | 20 (90.9) | 0.29*** |
| Deceased | 11 (18.3) | 9 (23.7) | 2 (9.1) |  |
| **Donor gender (%)** |  |  |  |  |
| Female | 32 (53.3) | 18 (47.4) | 14 (63.6) | 0.22** |
| Male | 28 (46.7) | 20 (52.6) | 8 (36.4) |  |

CMV: cytomegalovirus, SD: standard deviation, \*: t- student test, \*\*: Chi-square test, \*\*\*: Fisher's exact test, HTA hypertensive arterial.

**Table 1b.** Comparison of patient’s characteristics in those with CMV reactivation and those without, after kidney transplantation.

| Characteristic | Total(N=60) | With CMV reactivation(N=38) | No CMV reactivation(N=22) | p- value |
| --- | --- | --- | --- | --- |
| **Blood transfusion history** |  |  |  |  |
| No | 34 (58.6) | 21 (56.8) | 13 (61.9) | 0.7** |
| Yes | 24 (41.4) | 16 (43.2) | 8 (38.1) |  |
| **Group compatibility (%)** |  |  |  |  |
| Identical | 42 (70) | 27 (71.1) | 15 (68.2) | 0.81** |
| Different but compatible | 18 (30) | 11(28.9) | 7 (31.8) |  |
| **HLA-AB mismatch (%)** |  |  |  |  |
| 1 – 4 | 52 (88.1) | 33 (89.2) | 19 (86.4) | 1*** |
| 0 | 7 (11.9) | 4 (10.8) | 3 (13.6) |  |
| **HLA-DR mismatch (%)** |  |  |  |  |
| 1 – 2 | 44 (75.9) | 30 (83.3) | 14 (63.6) | 0.09** |
| 0 | 14 (24.1) | 6 (16.7) | 8 (36.4) |  |
| **Induction therapy (%)** |  |  |  |  |
| Thymoglobuline | 50 (83.3) | 32 (84.2) | 18(81.8) | 1*** |
| Basiliximab | 10 (16.7) | 6 (15.8) | 4 (18.2) |  |
| **Anti-calcineurin treatment (%)** |  |  |  |  |
| Cyclosporine | 1 (1.7) | 1 (2.6) | 0 | 1*** |
| Tacrolimus | 49 (83.1) | 31 (81.6) | 18 (85.7) |  |
| Cyclosporine + Tacrolimus | 9 (15.3) | 6 (15.8) | 3 (14.3) |  |
| **Traitement Inhibition cellular multiplication treatment (%)** |  |  |  |  |
| Mofetil Mycophenolat | 47 (78.3) | 31 (81.6) | 16 (72.7) | 0.34*** |
| Mycophenolat acid (Myfortic) | 1 (1.7) | 0 | 1 (4.5) |  |
| MMF follow -up by Myfortic | 10 (16.7) | 5 (13.2) | 5 (22.7) |  |
| MMF follow-up by Azathioprine | 2 (3.3) | 2 (5.3) | 0 |  |
| **Duration of primary CMV prophylaxis (days), mean ± SD** | 125 ± 45 | 119 ± 48 | 136 ± 39 | 0.16* |
| **Patients receiving antiviral prophylaxis (%)** |  |  |  |  |
| Valaciclovir | 48 (80) | 32 (84.2) | 16 (72.7) | 0.33*** |
| Valganciclovir | 12 (20) | 6 (15.8) | 6 (27.3) |  |
| **Opportunistic infections (%)** |  |  |  |  |
| No | 25 (43.1) | 13 (34.2) | 12 (60) | 0.06** |
| Yes | 33 (56.9) | 25 (65.8) | 8 (40) |  |

HLA: human leukocytes antigens, ATG: anti-thymocyte globulin, MMF: mycophenolate mofetil, CMV: cytomegalovirus, SD: standard deviation, \*: t- student test, \*\*: Chi-square test, \*\*\*: Fisher's exact test.

**Fig 2.**
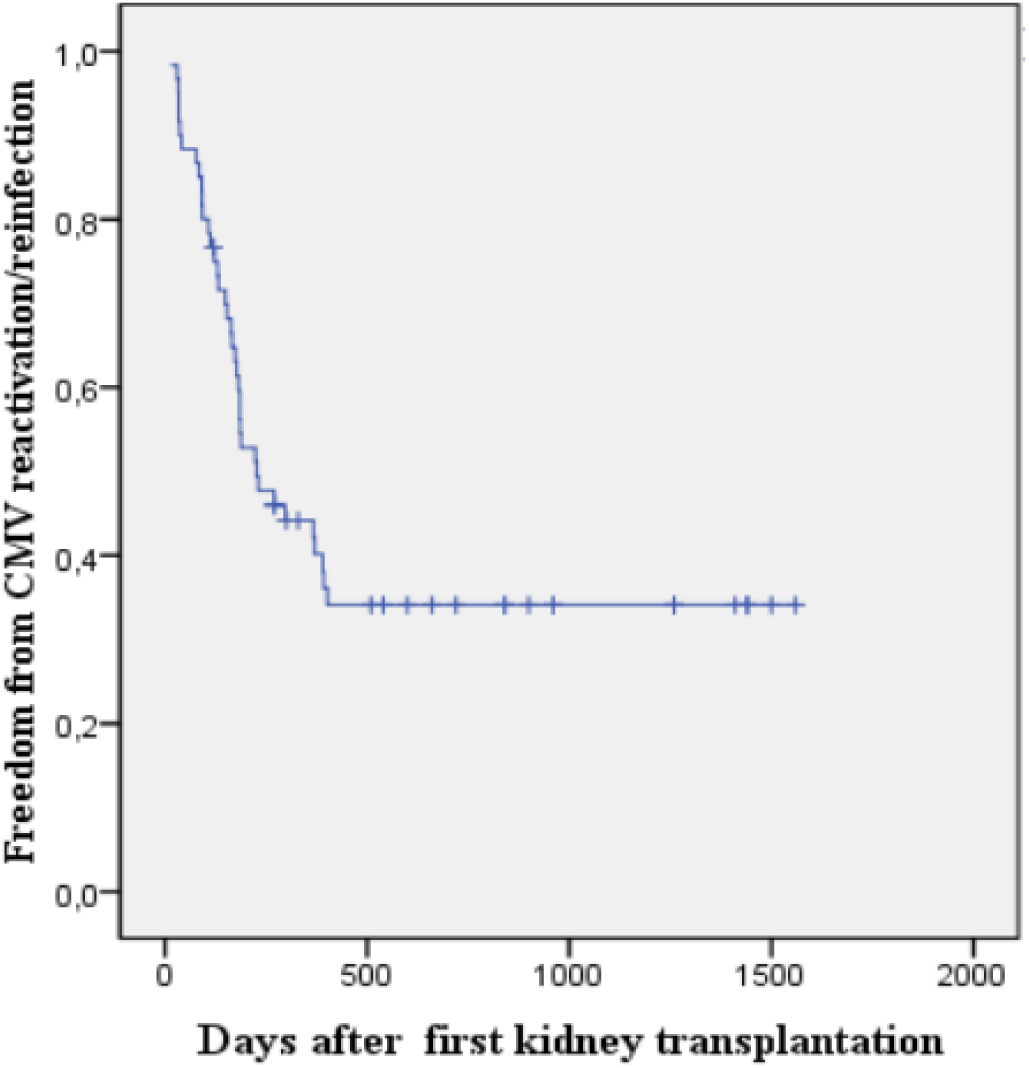
Median time of reactivation/reinfection in seropositive kidney recipients after first kidney transplantation using Kaplan Meier. The time course of CMV reactivation was measured after kidney transplantation during the follow-up period. We illustrated the follow -up time on the axis for all seropositive patients (with experienced CMV replication and without).

In the context of CMV viraemia, thirty out of the reactivated patients (78.9%) had at least one episode of quantitative viral load (mean = 2.4 ± 1.19 log IU/ml) with [1.49 to 6.79] log IU/ml. Eight (21.1%) patients had developed one episode of detectable viraemia below the quantification threshold (<1.49log_10_IU/ml).

Significantly lower period of viral replication after transplantation was observed among the transplant patients with CMV quantifiable viral load compared to patients with only detectable viraemia (141.5 ± 96.9 vs. 294.1 ± 112.4 days, *P* < 0.001). Furthermore, A reactivation at the time of prophylactic treatment was significantly more frequent in patients who experienced a quantifiable viral load, compared to patients with only detectable viraemia (14 (47%) vs. 0 (0%), *P* = 0.017). Nevertheless, reactivation with quantifiable viral load occurred earlier after the end of prophylaxis in patients who presented an important level viral load (78.1 ± 70.3 vs. 166.8 ± 118.3 days, *P* = 0.06). Interestingly, no differences were observed in the total average of leukocyte, neutrophils polynuclear or lymphocytes counts at diagnosis between patients with or without quantifiable viraemia. Additional CMV viraemia related outcomes are shown in Table 2.

**Table 2.** Comparison of patient’s characteristics in those who developed quantitative detectable viraemia versus in those who did not.

| Characteristic | Total (N=38) | With detectable viraemia(N=8) | With quantitative viraemia (N=30) | p- value |
| --- | --- | --- | --- | --- |
| Age, mean and SD, years | 36.6 ± 13.19 | 40.3 ± 18.53 | 35.6 ± 11.58 | 0.37* |
| Sex (%) |  |  |  |  |
| Female | 15 (39.5) | 4 (50) | 11 (36.7) | 0.69** |
| Male | 23 (60.5) | 4 (50) | 19 (63.3) |  |
| Source of donor (%) |  |  |  |  |
| Living | 29 (76.3) | 6 (75) | 23 (76.7) | 1** |
| Deceased | 9 (23.7) | 2 (25) | 7 (23.3) |  |
| Blood transfusion history (%) |  |  |  |  |
| No | 21 (56.8) | 2 (25) | 19 (65.5) | 0.055** |
| Yes | 16 (43.2) | 6 (75) | 10 (34.5) |  |
| HLA- AB mismatch (%) |  |  |  |  |
| 1 -4 | 33 (89.2) | 8 | 25 (86.2) | 0.56** |
| 0 | 4 (10.8) | 0 | 4 (13.8) |  |
| HLA -DR mismatch (%) |  |  |  |  |
| 1 -2 | 30 (83.3) | 7 (87.5) | 23 (82.1) | 1** |
| 0 | 6 (16.7) | 1 (12.5) | 5 (17.9) |  |
| Duration of prophylactic treatment (days), mean ± SD | 118.7 ± 48.13 | 107.3 ± 28.43 | 121.7 ± 52.11 | 0.46* |
| Time of active replication after transplantation (days), mean ± SD | 173.6 ± 117.18 | 294.1 ± 112.6 | 141.5 ± 96.9 | < 0.001* |
| Active replication at prophylactic period treatment (%) |  |  |  |  |
| No | 24 (63.2) | 8 | 16 (53) | 0.017** |
| Yes | 14 (36.8) | 0 | 14 (47) |  |
| Time of active replication after prophylactic treatment (days), mean ± SD | 95.8 ± 86.25 | 166.7 ± 118.33 | 78.1 ± 70.34 | 0.064* |
| Mean leukocytes count at diagnosis ± SD (/ml) | 6605 ± 3012 | 5278 ± 1550 | 6880 ± 3183 | 0.24* |
| Mean polynuclear neutrophils counts at diagnosis ± SD (/ml) | 5136 ± 2911 | 3713 ± 1708 | 5430 ± 3040 | 0.19* |
| Mean lymphocytes count at diagnosis ± SD (/ml) | 845 ± 518 | 1090 ± 382 | 795 ± 533 | 0.21* |
| Intercurrent infections |  |  |  |  |
| No | 28 (73.7) | 7 (87.5) | 21 (70) | 0.65* |
| Yes | 10 (26.3) | 1 (12.5) | 9 (30) |  |

SD: standard deviation, HLA: human leukocytes antigens, \*: t- student test, \*\*: Fisher's exact test .

CMV disease occurred in eight (13%) of the transplanted population. Among patients with quantifiable viraemia, CMV infection progressed to CMV syndrome or disease in 8/30 (27%) and 22/30 (73%) of reactivated patients were with asymptomatic viraemia.

The incidence rate of symptomatic reactivation was six patients per 100 person-year. Most symptomatic recipients developed CMV syndrome or disease within the first five months from solid organ transplantation (7/8, 87.5%). The CMV syndrome was diagnosed in 2/8 (25%) of symptomatic patients and 6/8 (75%) patients experienced CMV disease. Two patients with syndrome infection presented a low-grade fever, mild lymphocytopenia and low increased level of alanine aminotransferase (ALT). Among patients with CMV disease, diarrhea, leucopenia, lymphopenia and low impairment in renal function were more frequently observed. One patient had both diarrheas, vomiting and severe abdominal pain without confirmed by tissue-invasive.

Interestingly for demographic characteristics, younger patients were significantly associated with CMV disease reactivation (28.8 ± 5.12 vs. 38.1 ± 12.34, *P* = 0.007) (Table 3a). Despite using the dose of all immunosuppressive drugs as thymoglobuline induction, mycophenolate mofetil, tacrolimus or cyclosporine, no effect was demonstrated on viral disease reactivation.

**Table 3a.** Comparison of patient’s characteristics in those with symptomatic CMV reactivation and in those with asymptomatic reactivation.

| Characteristic | Total (N=30) | With symptomatic reactivation (n=8) | With asymptomatic reactivation (n=22) | p- value |
| --- | --- | --- | --- | --- |
| **Recipient Age at transplantation in years(± SD)** | 35.6 ± 11.58 | 28.8 ± 5.12 | 38.1 ± 12.34 | <u>0.007</u>* |
| **Donor age at transplantation in years (± SD)** | 38.5 ± 14.22 | 36.8 ± 14.87 | 39.1 ± 14.28 | 0.69* |
| **Donor source** |  |  |  |  |
| Living | 23 (76.7) | 5 (62.5) | 18 (81.8) | 0.34* |
| Deceased | 7 (23.3) | 3 (37.5) | 4 (18.2) |  |
| **HLA-AB mismatch (%)** |  |  |  |  |
| 1 -4 | 25 (86.2) | 6 (85.7) | 19 (86.4) | 1** |
| 0 | 4 (13.8) | 1 (14.3) | 3 (13.6) |  |
| **HLA -DR (%)** |  |  |  |  |
| 1 -2 | 23 (82.1) | 5 (71.4) | 18 (85.7) | 0.57** |
| 0 | 5 (17.9) | 2 (28.6) | 3 (14.3) |  |
| **Induction treatment (%)** |  |  |  |  |
| Thymoglobulin | 26 (86.7) | 7(87.5) | 19(86.4) | 1** |
| Basiliximab | 4 (13.3) | 1(12.5) | 3 (13.6) |  |
| **Average creatinine level(mg/l) mean at 3 months of monitoring (± SD)** | 11.2 ± 4.16 | 11.2 ± 4.65 | 11.2 ± 4.07 | 0.99* |
| **Average creatine clearance rate (ml/min/1.73m<sup>2</sup>) at 3 months of monitoring (± SD)** | 78.7 ± 23.13 | 81.3 ± 28.21 | 77.7 ± 21.53 | 0.71* |
| **Mean leukocyte counts(/ml) at 3 months of monitoring (± SD)** | 6240 ± 3007 | 5131 ± 1076 | 6706 ± 3441 | 0.08* |
| **Mean polynuclear neutrophil counts (/ml) at 3 months of monitoring (± SD)** | 4784 ± 2744 | 3602 ± 1074 | 5282 ± 3090 | 0.047* |
| **Mean lymphocytes counts (/ml) at 3 months of monitoring (± SD)** | 858 ± 476 | 944 ± 474 | 822 ± 485 | 0.55* |

CMV: cytomegalovirus, SD: standard deviation, HLA: human leukocyte antigen, \*: t- student, \*\*: Exact Fisher test,

**Table 3b.** Comparison of patient’s characteristics in those with symptomatic CMV reactivation and in those with asymptomatic reactivation.

| Characteristic | Total (N= 30) | With symptomatic reactivation (n= 8) | With asymptomatic reactivation (n= 22) | p- value |
| --- | --- | --- | --- | --- |
| **Average creatinine level(mg/l) mean at 6 months of monitoring (± SD)** | 11.0 ± 3.88 | 12.5 ± 5.77 | 10.4 ± 2.79 | 0.34* |
| **Average creatine clearance rate (ml/min/1.73m<sup>2</sup>) at 6 months of monitoring (± SD)** | 83.1 ± 27.47 | 80.2 ± 40.44 | 84.2 ± 21.56 | 0.79* |
| **Mean leukocyte counts(/ml) at 6 months of monitoring (± SD)** | 5820.3 ± 1748.18 | 5540.0 ± 2677.63 | 5938.4 ± 1255.61 | 0.69* |
| **Mean polynuclear neutrophil counts (/ml) at 6 months of monitoring (± SD)** | 3902.7 ± 1396.44 | 3795.3 ± 2264.46 | 3948.0 ± 902.94 | 0.86* |
| **Mean lymphocytes counts (/ml) at 6 months of monitoring (± SD)** | 1099.8 ± 669.48 | 1045.3 ± 688.12 | 1122.8 ± 679.25 | 0.79* |
| **Antiviral prophylaxis (%)** |  |  |  |  |
| Valaciclovir | 26 (86.7) | 6 (75) | 20 (90.9) | 0.39** |
| Valganciclovir | 3 (10) | 1(12.5) | 2 (9.1) |  |
| Valaciclovir follow-up Valganciclovir | 1 (3.3) | 1 (12.5) | 0 |  |
| **Duration of prophylactic treatment (days) (± SD)** | 124.9 ± 47.04 | 146.2 ± 34.87 | 138.5 ± 35.89 | 0.60* |
| **Mean CMV viral load at diagnosis (logUI/ml)** | 2.3 ± 1.19 | 2.9 ± 1.91 | 2.1 ± 0.74 | 0.29 * |
| **Mean lymphocytes counts at diagnosis (/ml)** | 795 ± 533 | 674 ± 424 | 841 ± 572 | 0.46 * |
| **Duration of appearance reactivation after kidney transplantation (days)** | 132.9 ± 84.10 | 140.1 ± 109.20 | 130.3 ± 75.93 | 0.78* |

CMV: cytomegalovirus, SD: standard deviation, HLA: human leukocyte antigen, \*: t- student, \*\*: Exact Fisher test.

**Table 3c.**
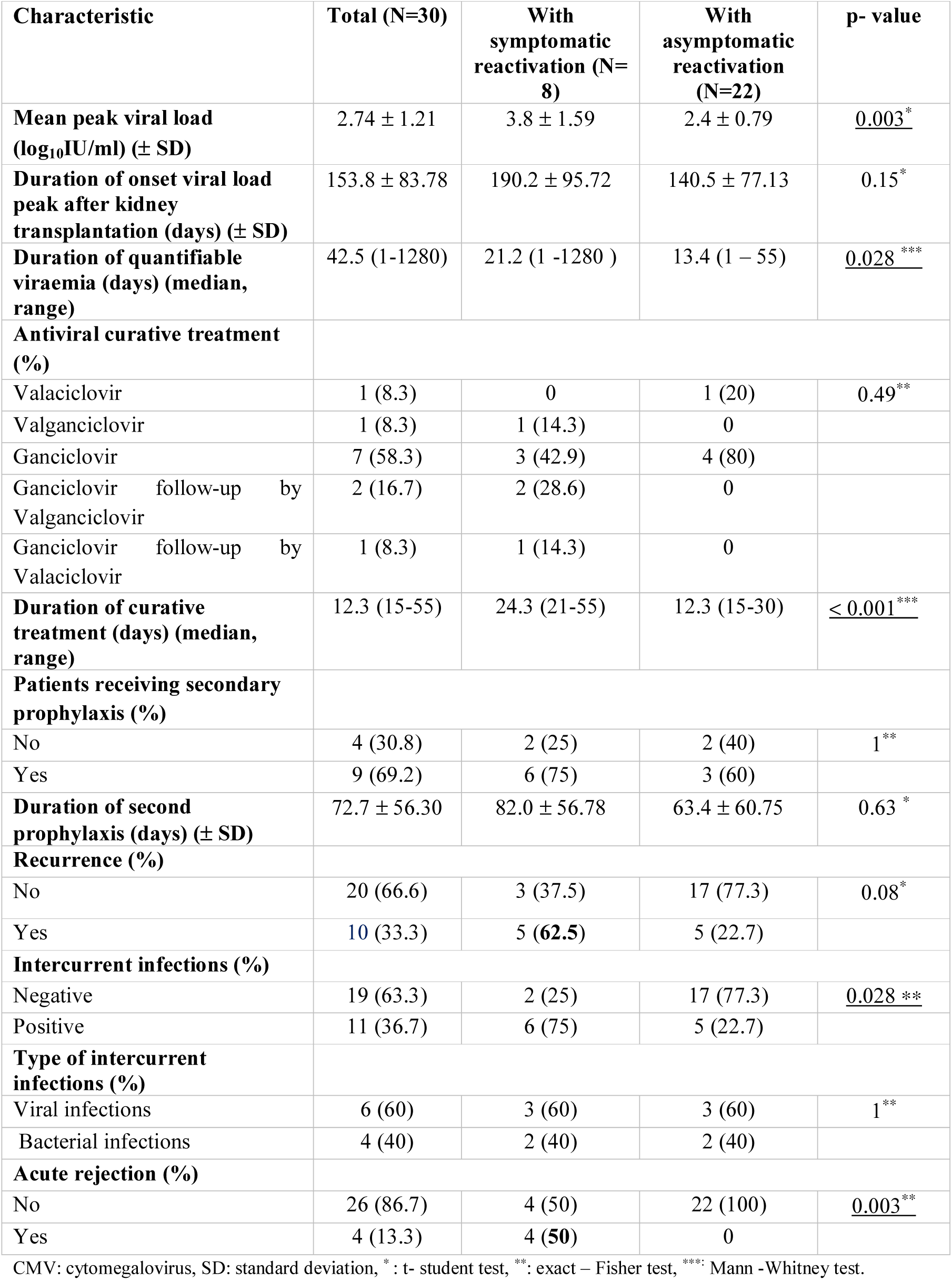
Comparison of patient’s characteristics in those with symptomatic CMV reactivation and in those with asymptomatic reactivation.

Significant higher peak viral load was also observed for patients with CMV disease compared with those who remained symptoms free (3.8 ± 1.59 vs. 2.4 ± 0.79, *P* = 0.003). Furthermore, the median duration of viraemia appeared to be also statistically significant for this group 21.2 [1 to 570 days] vs. 13.4 [1 to 42 days], *P* = 0.028.

Symptomatic patients were treated with either intravenous ganciclovir alone or with relay by valacyclovir or valganciclovir. One patient who suffered from serious dysfunction transplant did not receive curative treatment and was treated only with reduction of immunosuppression (MMF). In addition, mycophenolate dose was temporally reduced for all symptomatic patients. On the other hand, five patients without CMV related symptoms but who showed a high viral load were also treated with intravenous ganciclovir and one patient received only oral valaciclovir (Table 3a, 3b). Seventeen asymptomatic patients with low viral load were monitored regularly by real-time PCR without interventions. The median duration of antiviral treatment was 12 [15 to 55] days. However, the duration of CMV therapy was significantly longer in the symptomatic patients than in the asymptomatic recipients (median period = 24.3 days ranging 21-55 vs. median = 12.3 days ranging 15 - 30, *P* < 0.001, respectively). Nine cases were suspected of antiviral resistance with persistence of viraemia after three weeks of treatment or increased viral load at time of curative treatment. After achieving viral load clearance, a secondary prophylaxis was given to nine patients who received antiviral therapy (Table 3b). Viral Infections with polyoma BK virus was more observed in this group. Likewise, most patients with symptomatic CMV reactivation showed significantly high proportion of intercurrent infections compared to asymptomatic patients (6/8 (75%) vs. 5/22 (23%), *P* = 0.028). Furthermore, acute graft rejection was statistically recorded in 4/8 (50% with *P* = 0.003, Table 3c). In the other hand, 6% of population had recurrent CMV and was seen more frequently among patients who presented syndrome or disease compared to patients without symptoms (62.5% vs 22.7%, *P* = 0.08).

The incidence of recurrence CMV was 10 (17%) of 60 patients. Recurrence of CMV infection occurred an average of 288.9 ± 206.4 days after transplantation, or appeared after first episode of reactivation at mean 136.2 ± 120.5 days. The duration of recurrence was at median 5.5 days (range 1-710 days). After discontinuation of antiviral therapy, recurrence was observed at mean time of 158.4 ± 139.7days. Among patients with recurrence, 3/10 (30%) had only detectable viraemia and seven of 10 patients (70 %) developed quantifiable viraemia which an average viral load at diagnosis was 2.3 ± 0.8 log_10_IU/ml. In addition, mean peak viral load of recurrence remained higher than in no recurrent patients (2.7 ± 0.7 log_10_IU/ml). In this study, the most recurrent patients had no clinical symptoms (8/10, 80%). But, two recipients (20%) with symptomatic CMV recurrence presented either with hepatitis, hematuria or diarrhea. All patients with higher viral load were treated with intravenous GCV and carefully monitored until viraemia subsided. Renal function (plasma creatinine and estimated glomerular filtration rate eGFR) and blood white cell counts (leukocyte, polynuclear neutrophils and lymphocyte) measured at 3 and 6 months of follow-up in patients with recurrent infection were similar (Table 4). For baseline characteristics and outcomes of recurrence, no significative differences were observed between patients with or without recurrent CMV infections (Table 4).

**Table 4.** Comparison of patient’s characteristics with and those without CMV recurrence.

| Characteristic | Total<br>(N = 30) | With CMV<br>recurrence<br>(N= 10) | Without CMV<br>recurrence<br>(N = 20) | p-value |
| --- | --- | --- | --- | --- |
| Age at transplantation, mean years (±SD) | 35.6 ± 11.58 | 31.2 ± 12.44 | 37.8 ± 10.78 | 0.14* |
| Sex (%) |  |  |  |  |
| Female | 11(36.7) | 3 (30) | 8 (40) | 0.70** |
| Male | 19 (63.3) | 7 (70) | 12 (60) |  |
| Plasma creatinine at 3 months of follow-up | 11.3± 4.16 | 12.3 ± 4.13 | 10.8 ± 4.19 | 0.38* |
| Clearance creatine at 3 months of follow-up | 78.8 ± 23.13 | 75.7 ± 27.93 | 80.2 ± 21.19 | 0.64* |
| Leukocytes counts at 3 months of follow-up (/ml) | 6240 ± 3008 | 5274 ± 2849 | 6723 ± 3046 | 0.25* |
| Polynuclear neutrophils counts (/ml) at 3 months of follow-up | 4784.6 ± 2711.7 | 3866.8± 2616.7 | 5243.5 ± 2762.6 | 0.23* |
| Lymphocytes counts (/ml) at 3 months of follow-up | 858 ± 476 | 920 ± 496 | 827 ± 477 | 0.64* |
| Plasma creatinine at 6 months of follow-up (ml/min/1.73m<sup>2</sup>) | 11.0 ± 3.9 | 12.9 ± 4.4 | 9.9 ± 3.2 | 0.05* |
| Clearance creatine at 6 months of follow-up (ml/min/1.73m<sup>2</sup>) | 83.1 ± 27.5 | 73.8 ± 30.01 | 88.2 ± 25.3 | 0.19* |
| Leukocytes counts at 6 months of follow-up (/ml) | 5820 ± 1748 | 5971 ± 2274 | 5745 ± 1490 | 0.76* |
| Polynuclear neutrophils counts (/ml) at 6 months of follow-up | 3902 ± 1396 | 4054 ± 1953 | 3826 ± 1081 | 0.75* |
| Lymphocytes counts (/ml) at 6 months of follow-up | 1099 ± 669 | 1209 ± 990 | 1045 ± 463 | 0.65* |
| HLA- AB mismatch (%) |  |  |  |  |
| No | 4 (13.8) | 1(11.1) | 3 (15) | 1** |
| Yes | 25 (86.2) | 8 (88.9) | 17 (85) |  |
| HLA-DR mismatch (%) |  |  |  |  |
| No | 5 (17.9) | 2 (25) | 3 (15) | 0.61** |
| Yes | 23 (82.1) | 6 (75) | 17 (85) |  |
| Induction treatment (%) |  |  |  |  |
| Thymoglobulin | 26 (86.7) | 8 (80) | 18 (90) | 0.58** |
| Basiliximab | 4 (13.3) | 2 (20) | 2 (10) |  |
| Anti-calcineurin treatment (%) |  |  |  |  |
| Cyclosporine | 2 (6.7) | 2 (20) | 0 | 0.09* |
| Tacrolimus | 23 (76.7) | 6 (60) | 17 (85) |  |
| Cyclosporine follow-up by Tacrolimus | 5 (16.7) | 2 (20) | 3 (15) |  |

CMV: cytomegalovirus, SD: standard deviation, \*: t- student test, \*\*: exact – Fisher test.

### Univariate analysis regression logistic

In the present study, first univariate analysis was performed in order to determine risk factors related to reactivation and recurrence. In this model, univariate analysis included factors that were associated with quantitative viraemia and symptomatic reactivation. We specifically viral factors that influenced symptomatic reactivation (Table 5). Following renal transplantation, peak viral load has significantly predicted the risk of symptomatic CMV reactivation [OR 3.39 (95% IC1.21, 9.53), *(P* = 0.02)], whereas time of active replication was related to occurrence with quantitative viraemia [OR 0.98 (95% IC 0.98, 0.99), (*P*= 0.004)]. The risk associated with viral load at diagnosis, antiviral prophylactic treatment (generally with valaciclovir) and duration of primary prophylaxis were not associated with symptomatic CMV reactivation (*P*= 0.14, *P* = 0.18 and *P* = 0.06, respectively).

**Tableau 5.** Analysis of risk factors of reactivation with quantitative viraemia, clinical symptoms and recurrence CMV infection in kidney seropositive patients (D+/R+).

| Variable | CMV reactivation T(n=38) |  | Reactivation with quantitative viraemia (n=30) |  | Reactivation with clinical symptomatic CMV(n=8) |  | CMV recurrence (n=10) |  |
| --- | --- | --- | --- | --- | --- | --- | --- | --- |
|  | Odds ratio (95% CI) | P-value* | Odds ratio (95% CI) | P-value* | Odds ratio (95% CI) | P-value* | Odds ratio (95% CI) | P- value* |
| Age of recipients at transplantation (years) | 0.96 (0.92,1.00) | 0.08 | 0.97 (0.92, 1.03) | 0.36 | 0.92(0.83, 1.01) | 0.07 | 0.95 (0.88,1.02) | 0.15 |
| Donor age (years) | 0.98(0.95,1.02) | 0.37 | 1.00 (0.95, 1.06) | 0.96 | 0.99 (0.93, 1.05) | 0.70 | 1.02 (0.96, 1.07) | 0.60 |
| Deceased donor | 3.10(0.61,15.91 ) | 0.17 | 0.91 (0.15, 5.58) | 0.92 | 2.7 (0.45,16.26) | 0.28 | 1.71 (0.30, 9.77) | 0.54 |
| Number of HLA-DR mismatch | 2.86(0.83,9.82) | 0.09 | 0.66 (0.07, 6.61) | 0.72 | 0.42 (0.05, 3.22) | 0.40 | 0.53 (0.07, 3.98) | 0.54 |
| Blood transfusion history | 0.81(0.27, 2.41) | 0.70 | 5.7 (0.97, 33.60) | 0.05 | 3.56 (0.61, 20.81) | 0.16 | 0.93(0.18, 4.90) | 0.93 |
| Time of active replication after transplantation (days) | NA | NA | 0.99 (0.98, 0.99) | <u>0.004</u> | 1 (0.99, 1.01) | 0.76 | NA | NA |
| Viral load at diagnosis (log<sub>10</sub> IU/ml) | NA | NA | NA | NA | 1.71 (0.83, 3.50) | 0.14 | NA | NA |
| Mean Peak viral load (log<sub>10</sub> IU/ml) | NA | NA | NA | NA | 3.39 (1.21, 9.53) | <u>0.02</u> | 1.65 (0.83, 3.30) | 0.16 |
| Antiviral prophylaxis | 0.50 (0.14,1.8) | 0.29 | 0.46 (0.07, 3.13) | 0.43 | 3.38 (0.58, 19.60) | 0.18 | 6.6 (0.69, 63.25) | 0.10 |
| Length of primary prophylaxis (days) | 0.99 (0.98,1.00) | 0.16 | 1.01 (0.99, 1.02) | 0.45 | 1.03 (0.99, 1.06) | 0.06 | 0.99 (0.97, 1.01) | 0.53 |

CMV: Cytomegalovirus, D+: seropositive donor, R+: seropositive recipient, HLA: human leukocytes antigens, IU: international unit, NA: not applicable.
\*p-values were calculated using univariate analysis regression logistic.

## Discussion

We found that virus replication after renal transplantation is frequent in seropositive recipients. Despite prophylaxis, 63% of first episodes reactivation was observed in this group. This finding was in accordance with data of recent literature, 43 to 61.9% CMV seropositive transplant patients had experienced virus reactivation or re-infection by detection of viral DNA [18, 29]. One study [30] demonstrated that transplanted organ of seropositive donor was associated with 49% infection rates in the recipients with lack of prophylactic treatment. The other study [15] has reported that if the donor was CMV seropositive, the risk of CMV reactivation was found to be more than doubled within the first-year post-transplantation.

We postulate that this high rate could indicate the role of seropositive donor who can play through the recipient’s exposure to new virus strains. A number of previous studies had suggested that seropositive patients were more likely to be re-infected with new strains than to have active replication of the latent virus [31]. Otherwise, CMV reactivation could also be caused by contact with immunocompetent infected individuals after post transplantation where seroprevalence of CMV reached 96.7% in our population [32].

Nevertheless, others studies reported that kidney transplant recipients were commonly exposed to the risk of reactivation as they received a high immunosuppression treatment during the first three months of follow-up [25, 33]. However, in our study, induction therapy with lymphocyte-deleting agents as ATG (thymoglobulin) did not show a significant difference compared to basiliximab. Moreover, our data demonstrated that reactivated patients receiving triple maintenance therapy such as tacrolimus, mycophenolate and steroids did not reach statistical results.

CMV replication has been known as dynamic process in the human host. Therefore, the management of CMV levels relies on quantitative analysis of CMV markers. In the current era, several tests were proposed for detecting and monitoring CMV replication after solid organ transplantation. However, quantitative real -time PCR is now the standard assay for measuring viral load with increased sensitivity [16, 34]. In our study, 79 % of reactivated patients had shown quantitative viral load with a limit detection threshold of 1.49 log_10_ IU/ml. The incidence of symptomatic viraemia (13%) among our transplant population was similar with previous study that included only seropositive kidney transplants [33]. However, our result seemed to be lower in comparison with Madi’s data which 68.5% of most kidney recipients had symptomatic infection [35]. We postulate that the different variations observed between our results and those of others studies could be attributed either to CMV preventions strategies (universal prophylaxis versus preemptive treatment), or to antiviral drugs used (valaciclovir versus valganciclovir or ganciclovir), and longer duration of prophylaxis treatment (six versus three months) for R+ recipients. However, the use of prophylaxis treatment for D+/R+ group did not spare a subset of patients to be exposed to CMV disease.

Additionally, among the group of symptomatic CMV infection, the mean peak viral load was greater than in patients with asymptomatic viraemia. This finding was corroborated by the results of systematic review and meta-analysis [36] which indicated that mean viral load remained significantly higher in patients with CMV disease [OR 9.3 (95% CI, 4.6-19.3)]. Furthermore, these patients had substantially prolonged viraemia (average 21 versus 13 days, *P*= 0.028) to reach viral clearance compared to those with symptoms free. This finding supported previous study [37], which revealed that high initial viral load was associated with prolonged time to CMV clearance. According to several studies, the high viral load and rapid increase in plasma were associated with the occurrence of invasive CMV disease [1, 35, 38].

Advanced age was also considered as the main risk factor of development of symptomatic infection [9, 39, 40]. However, in our cohort, we found that the young patients were more likely to have symptomatic viral reactivation (28.75 ± 5.12 vs. 38.09 ± 12.34, p **<** 0.007). Several authors had suggested that reactivation was more frequent in elderly patients because they had a weakened immune system exacerbated by immunosuppressive treatment [18, 41]. Even if our population was composed mainly of young patients, the frequency of observations related to clinical manifestations does not exclude the possibility of interference from immunological factors. Indeed, one study (42) demonstrated through the QuantiFERON-CMV test regardless of age that patients with low levels of anti-CMV immune cells were more likely to experience intensive viraemia associated with severe clinical manifestations. Reasonably, it was impossible for us to explain this situation for our patients due to the lack of data on immunological parameters such as measurement of specific CMV-antibodies titers and T-cell lymphocytes.

Regards to clinical consequences of intensity viraemia, gastrointestinal manifestations were the most common of CMV disease reported in (4/8) 50% of symptomatic patients associated to laboratory abnormalities such as leucopenia, lymphopenia or elevated transaminases, and only two recipients (25%) developed CMV syndrome. These manifestations were observed before discontinuing treatment and without confirmation by invasive diagnosis. As opposed to previous report, fever was considered the most prevalent manifestation in such patients [43], however, a largest cohort of kidney transplant recipients study including 1129 patients from Germany and Finland revealed that gastrointestinal tract infections were detected in 46% and fever in 27% of patients who exhibited symptomatic reactivation [23]. Despite the use of prophylactic treatment was not routinely applied to CMV-kidney seropositive patients, the findings of this cohort study were almost similar to those of our present study which prophylaxis was extended to 6 months in our group of renal transplants.

However, in the current study, even with a significant high viral load measured in some cases, the clinical symptoms were not observed. This result was in accordance with previous study [35]. The author of this report indicated that 13 patients without CMV -related clinical consequences showed a high viral load. These discrepancies may be explained by different factors such as the presence of re-stimulated CMV specific cellular immunity in the host, limited dissemination of virus, source of reactivated CMV strains and possibly concomitant reactivation of several strains. Those patients who developed high-level viral reactivation without evidence of organ invasive disease may be exposed to indirect effects of CMV. Therefore, it is very important to implement a rigorous viral load monitoring throughout the post-transplant follow-up [44].

For kidney transplant recipients D+/R+, the use of pre-emptive treatment as preventive strategies has led to the definition of optimal viral load thresholds in order to initiate treatment or even predict the occurrence of disease or graft dysfunction. Thus, the majority of studies have explored several tests of real-time PCR in order to determine the optimal viral thresholds expressed in different units. In our context, we have also used quantitative PCR for detecting CMV DNA in plasma according to recommendations of international guideline [14]. But, we did not establish previously viral load thresholds for defining patients at high risk of CMV disease for that reason we should performed another specific study taking in account the baseline risk of the kidney seropositive patients for developing high viral loads. There is another significant point which we were unable to compare our results of viral loads with other data of different laboratory tests because there is still variability of measurement expression in units or copies notwithstanding use of international referenced to standard units defined by World Health of Organization like our study.

Interestingly, the time of therapeutic treatment was significantly twice longer than that of asymptomatic patients with only quantitative viraemia 24.3 [15 to 55] vs. 12.3 [15 to 30] days, p < 0.001. Otherwise, we found that the incidence of CMV disease during prophylaxis was 7/8 (87.5%) and 50% of symptomatic patients had CMV symptoms during the first three months of prophylactic treatment. Therefore, we also discovered very high rates of quantifiable viraemia and disease during prophylaxis while other studies did not. Recently, the transplantation society international CMV consensus group [16] suggested that valaciclovir can be used as a first line prophylactic drug for both D+/R- and D±/R+. Because, according to several randomized studies, the use of high dose valaciclovir significantly reduced the incidence of infection and even the severity of CMV disease [45 – 47]. Recent study [48] had demonstrated that valaciclovir was significantly effective in the prevention with reduction of the incidence of CMV infection (9.6% in valaciclovir group vs 67.6% in control group; p < 0.001). As result, our finding may be explained by the fact that all patients had received lower dose of valaciclovir varied from 4.5g to 6g /daily instead of 8g/daily as recommended dose for most of whom were given rATG induction therapy [47]. Another explanation would be proposed in the context of patient compliance that could affect the effectiveness of the treatment chosen. Indeed, in the absence of full coverage by the national health insurance system, patients would be more likely to reduce the number of antiviral treatments in order to optimize the cost of the antiviral medication despite the fact that it was associated to lower cost used in prophylaxis treatment [46, 49]. With regard to the use of valganciclovir as prophylactic treatment, we could not distinguish a significant difference between the symptomatic and symptom-free groups due to the limited number of patients receiving this type of antiviral drug. On the other hand, ganciclovir was the main antiviral agent for curative therapy. However, according to the Transplantation Society International Consensus group [16], nine cases were seemingly suspected to develop antiviral resistance during assessment. This observation cannot be valited to explain this phenomenon because the proportion of cases (9/30, 30%) observed was higher compared to other research data.

Three cases are highly suspected because they showed an extension of treatment beyond six weeks and one of these patients had shown a viraemia with a persistent high viral load during the first 570-days episode. The other six cases had reactivated during prophylactic treatment with valaciclovir in low dose. Such cases are considered to be weakly suspected for resistance. Thus, a second part of this research will be focused on the analysis of potential genetic mutations using the sequencing method.

Regarding the indirect CMV outcomes, previous studies demonstrated that CMV disease is an important risk factor for acute renal allograft rejection [50, 51]. In our study, we found that the four patients who developed transplant rejection had all a CMV disease (4 (50%) vs. 0%, *P* = 0.003). CMV disease can cause dysregulation in immune system. This imbalance in the immune system may increase the risk of transplant rejection. One study [52] showed that CMV disease can increase the risk of acute kidney transplant rejection, and factors controlling CMV infection, can reduce episode of acute rejection.

The rate was found higher than in the previously two published studies [47, 53] with 34% and 30%, respectively. Similarly, in another report, the high level of CMV viraemia was positively associated with acute rejection [OR= 3.27,95%IC = 1.08-9.4, *P*= 0.039] [54]. But in our study, the number of events was too small to assess a peak viral load effect on the occurrence of rejection episode.

Considering other infections than CMV, the occurrence of intercurrent infection was more frequent in those patients with symptomatic reactivation 6 (75%) vs. 5 (22.7%), *P*= 0.028. Our finding agreed with Blazquez-Navarro et al [54] who revealed a strong association between the reactivation of cytomegalovirus and BK virus regardless of the level of viral load measurement.

Another important finding was that our incidence of recurrence was (33%) in patients with quantifiable viraemia occurring during the first year after transplantation. Recently, few studies have reported similar results ranging from 30.5% to 36% [22 – 24]. However, the appearance of recurrence observed is relatively late in comparison with results of these previous studies. To further add to this, we were not able to show a clinical or virologic factor associated with increased risk of recurrence in our small study.

There are several significant limitations to consider. First, our study was conducted retrospectively with small sample size, which limited to performing more robust statistical tests to explore a several variables considered as risk factors. According to the recommendation is not available to include more than one variable per 10 events, we could not perform multivariate regression models for all factors with *P* less than 0.2 on univariate analysis. Second, we investigated in a single hospital center, which may limit a generalizability of our results. Third, we did not perform specific tests of CMV immune monitoring; it is possible that cases with high viral load without symptoms may have immune dysfunction during prophylaxis. Due to the fact that most of our patients were given valaciclovir as the basis for prophylactic treatment, it has been impossible to make a comparison with the data from the other studies.

Nevertheless, the important strengths of our study included representative population of typical kidney seropositive transplant recipient. All measurements of CMV DNAemia were performed in plasma by the same QNAT technique in the same laboratory and reported in international standards units. The same standardized protocol of immunosuppression and different strategies of treatment were adopted for our population of kidney recipients.

## Conclusion

There is increasing evidence, in the light of these results, that CMV reactivation remains serious problem for seropositive transplant recipients who were expected to be on antiviral prophylaxis. Patients with high level of viraemia may be at an increased risk of progression to CMV disease and adverse events such as acute rejection. In this sense, to support current prevention strategies, the use of immunological markers is essential associated with intensive viral load monitoring within the first year post-transplantation as well as the investigation of suspected antiviral resistance. Although not specifically examined in our study, there is also need to determine viral load thresholds at which preemptive therapy must be initiated for our patient’s populations.

## Data Availability

All relevant data are within the manuscript.

## Acknowledgments

The authors thank all patients who participated in the study and staff at the renal service in Ibn Sina hospital. Special thanks were also addressed to group members at central laboratory of virology for their technical assistance in performing the real-time PCR assays.

